# Dynamic dysregulation of IL-6 and genes functional in NETosis, complement and coagulation in severe COVID-19 illness

**DOI:** 10.1101/2020.10.13.20211425

**Authors:** Samanwoy Mukhopadhyay, Subrata Sinha, Saroj Kant Mohapatra

## Abstract

Comprehensive and unbiased re-analysis of published blood transcriptome data from patients of COVID-19 reveals significant up-regulation of the gene set functional in NETosis, but no evidence of general “cytokine storm”. In severe COVID-19 illness, there is significant up-regulation of complement and coagulation pathway, and negative correlation between NETosis and respiratory function (oxygen saturation). Interestingly, there is an early spike in the level of IL-6 gene expression in severe illness compared to moderate illness. With passing days post-onset, the level of IL-6 expression in severe illness approaches that in moderate illness. The data are consistent with IL-6 acting as a driver of NETosis in the early phase of severe COVID-19 illness, that results in a vicious cycle of NETosis-complement/coagulation-respiratory dysfunction. This has important consequence for timing of rational therapy with anti-IL-6 and NETosis inhibitors in severe COVID-19 illness.

## Introduction

COVID-19 disease caused by SARS-CoV-2 has rapidly become a center of intense scientific investigation, with emphasis on unraveling the biology for actionable knowledge. While the majority of the infected subjects are asymptomatic or mildly ill, a small percentage are severely ill with respiratory distress [1]. However, at present, it is difficult to predict with certainty the patients at high-risk for clinical severity and poor outcome, although multiple pathophysiological processes have been proposed, such as, cytokine storm [2, 3], coagulation and complement activation [4], neutrophil extracellular trap - NETosis [5]. These studies have also led to predictive biomarkers, such as, neutrophil: lymphocyte ratio [6, 7] and interleukin 6 (IL-6) expression [2, 4].

Insight into cytokine dysregulation has driven therapeutic advances, such as, anti-cytokine tocilizumab (IL-6 receptor antagonist) for severely ill patients of COVID-19 [8, 9]. Such treatment is premised on the induction of intra-pulmonary inflammation by SARS-Cov-2 infection that ultimately leads to severe local vascular dysfunction including micro-thrombosis, haemorrhage and pulmonary intravascular coagulopathy [10]. Therefore, it has been suggested to start tocilizumab early, in order to avoid mechanical ventilation [11], although the best timing for the treatment is still being investigated [12]. Therefore, it is important to understand the temporal and/or causal relationship of the cytokine up-regulation with coagulopathy and respiratory dysfunction.

The prothrombotic state (contributing to pulmonary dysfunction in COVID-19) is explained in terms of Neutrophil extracellular traps (NETs) that originate from decondensed chromatin of neutrophils that can trigger immunothrombosis. Critically ill patients of COVID-19 show significantly higher plasma levels of MPO-DNA complex, a marker of NETosis. Factors triggering NETs were significantly increased in COVID-19 and pulmonary autopsies confirmed NET-containing microthrombi with neutrophil-platelet infiltration. The authors concluded that NETs triggering immunothrombosis may partly explain the prothrombotic clinical presentations in COVID-19 [5].

In view of the pivotal role of cytokines (especially IL-6) and NETosis in biology of COVID-19 host response, we performed a deep and focussed investigation into IL-6, NETosis, complement and coagulation in published data from multiple patients of COVID-19 with varying illness severity. The primary goal was to dissect the transcriptomic dynamics of the functional modules and examine if there is a therapeutic window for drugs targeting specific pathophysiological mechanisms, such as IL-6 blockade or inhibition of NETosis.

## Results

Human transcriptomic data were extracted from published data sets of patients of COVID-19 from different tissues: whole blood (longitudinal sampling) [13], peripheral blood mononuclear cells (PBMC) [14], and lung tissue [15]. Whole blood was especially selected because it includes neutrophils that are directly responsible for formation of NETs.

### Up-regulation of NETosis in COVID-19

Targeted analysis was performed to study the extent of differential expression of two gene sets in whole blood: cytokine genes and NETosis genes. NETosis gene set was significantly up-regulated in whole blood of patients of COVID-19 [13]. Gene set enrichment analysis (permutation testing) revealed that the genes functional in NETosis were strongly up-regulated in the blood of COVID-19 patients compared to healthy subjects (Figure 1A). On the other hand, there was no evidence of broad up-regulation of cytokine gene set in the blood of the patients (Figure 1B).

**Figure 1:**
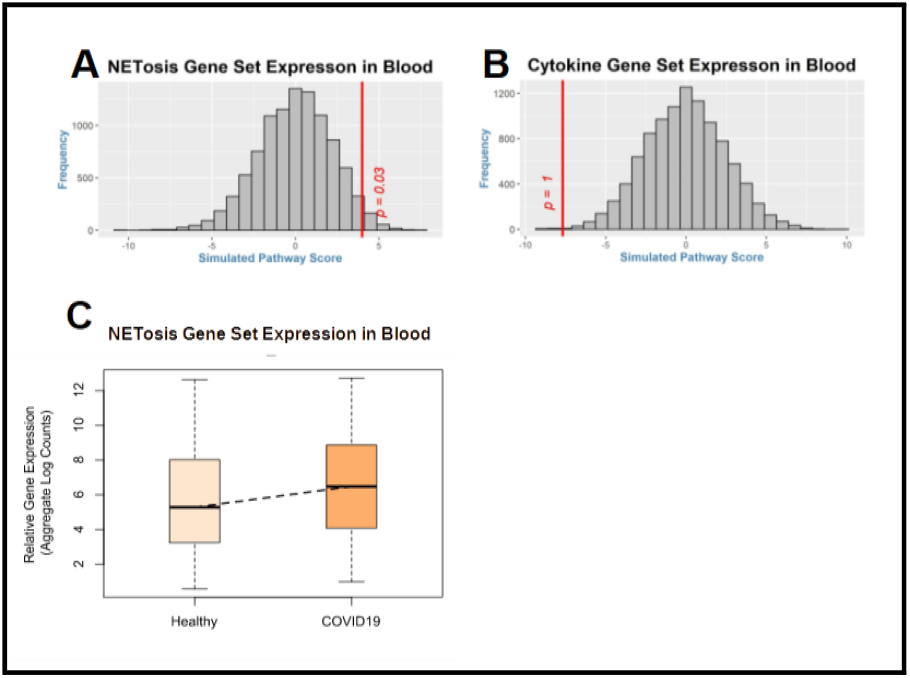
(A) For the gene set functional in NETosis, pathway score (red vertical line) was calculated by weighted averaging of t-statistic between control group (10 heathy subjects, single time point) and COVID-19 group (3 cases, multiple time-points). The histogram (gray bars) represents the null distribution of the pathway score calculated for each of the 10000 iterations of permuting the sample labels. The position of the red vertical line (observed pathway score) with respect to the histogram (null distribution) suggests that the pathway is significantly up-regulated in the blood of COVID-19 patients. As shown, there is significant (p = 0.03) up-regulation of the gene set functional in NETosis. (B) Calculation of the pathway score and the null-distribution of the cytokine gene set has been performed as mentioned for NETosis. As shown, there is no (p = 1) up-regulation of cytokine gene set in COVID-19. (C) For validation of NETosis, gene expression data of healthy control (n=6) and COVID-19 (n=7) were extracted from an independent cohort [14]. Box plot shows up-regulation of NETosis genes in COVID-19 cases compared to the control group. Gene expression data were extracted from [13] for panels A and B; and from [14] for panel C.

### Association of NETosis up-regulation with disease severity

As the heat map (Figure 2A) shows, the magnitude of up-regulation is greater in the severe COVID-19 (Case 1) compared to cases with moderate illness (Cases 2 and 3). In general, more genes functional in NETosis are up-regulated (red cells) in severe illness compared to moderate illness. The genes shown in Figure 2A are shown in the bar graphs in Figure 2B. Average level of expression of each gene across all time points was compared between moderate illness (blue bar) and severe illness (red bar). For each of these genes, level of expression is higher (and for many of these genes, the difference is statistically significant) in severe illness compared to moderate illness (Figure 2B).

**Figure 2A:**
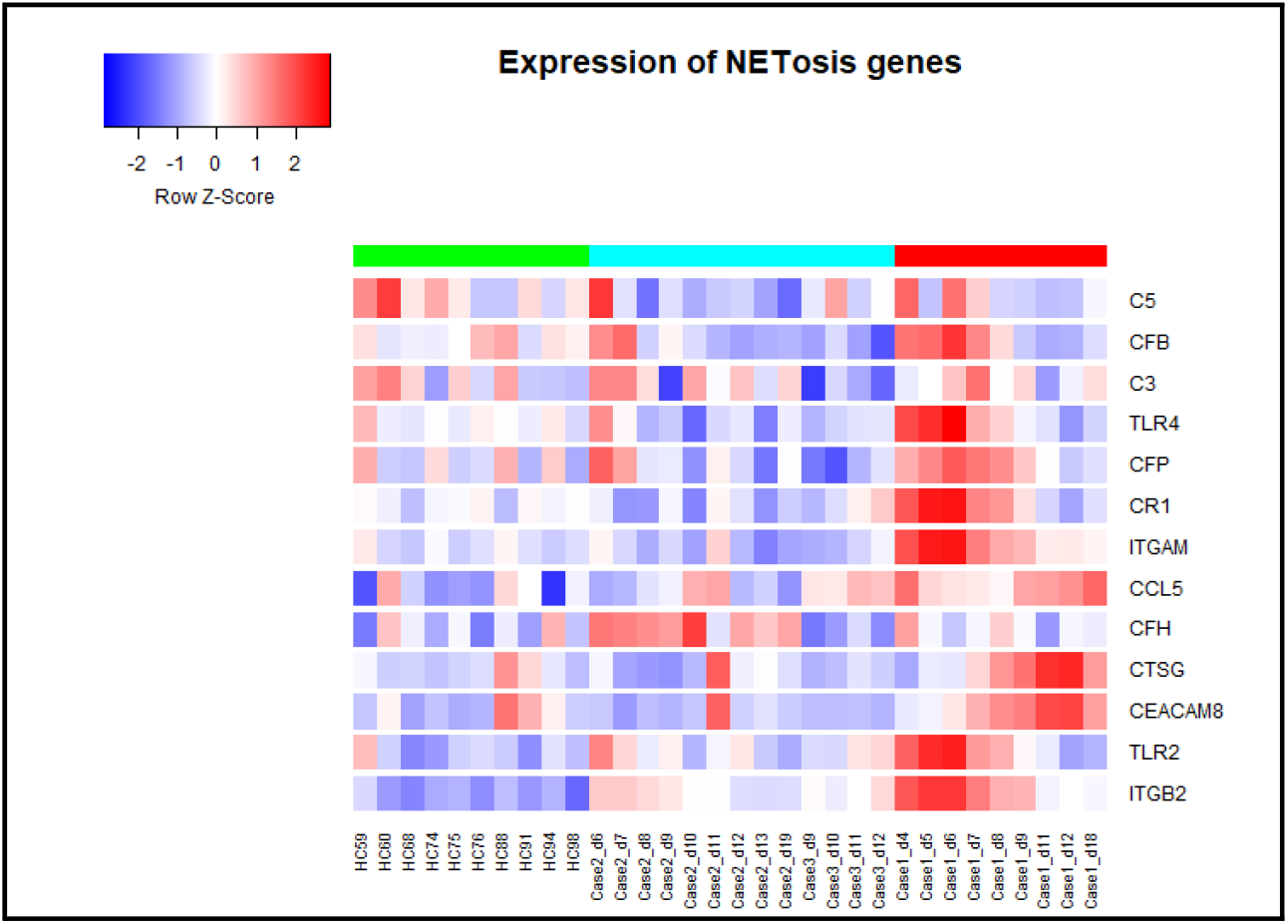
Column sidebar on the top marks samples from healthy control subjects as green, samples from moderate COVID-19 patients as cyan, and samples from severe COVID-19 patients as red. Each row represents a gene participating in the NETosis process. The color of the cell represents the level of expression, low as blue and high as red. Samples from 10 healthy control samples and 3 COVID-19 patients of varying illness severity (Case 1-severe, Cases 2, 3-moderate illness) at multiple time points are presented. The name of the sample includes case ID followed by the day post-onset of illness. Magnitude of up-regulation of NETosis genes is greater in the case with severe COVID-19 illness (as shown by the greater number of red cells for case 1 – severe illness). Gene expression data were extracted from [13].

**Figure 2B:**
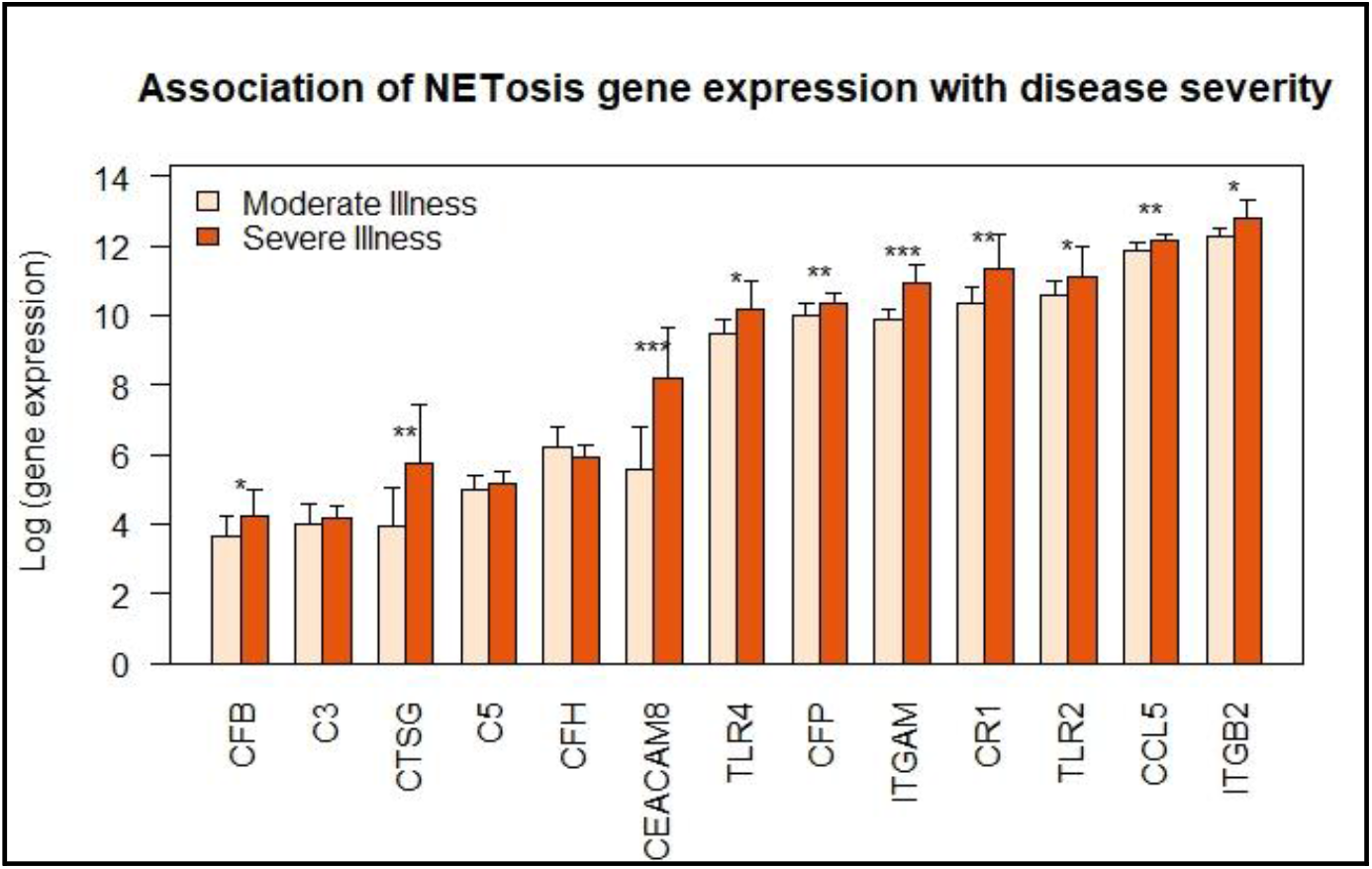
Each bar represents the mean log-expression of a NETosis gene across all-time samples from either moderate illness (2 cases) severe illness (single case). Error bar represents standard deviation of gene expression levels across all samples in that group of illness severity. For most of the genes there is a higher level of gene expression in severe illness compared to mild illness. Significance of up-regulation in severe illness was assessed by t-test and is indicated with an asterisk over the bars (* p < 0.1; ** p < 0.01; *** p < 0.001). Gene expression data were extracted from [13].

Further, level of gene expression was compared between severe and moderate illness for fixed time points (days post-onset of illness). For each of the four time points (Day 6, 7, 8 and 9), box plot was drawn to show up-regulation of NETosis in severe illness compared to moderate illness (Figure 2C). There is a statistically significant upward trend in gene expression from moderate illness to severe illness at each day post-onset, suggesting sustained up-regulation of genes functional in NETosis in severe illness.

**Figure 2C:**
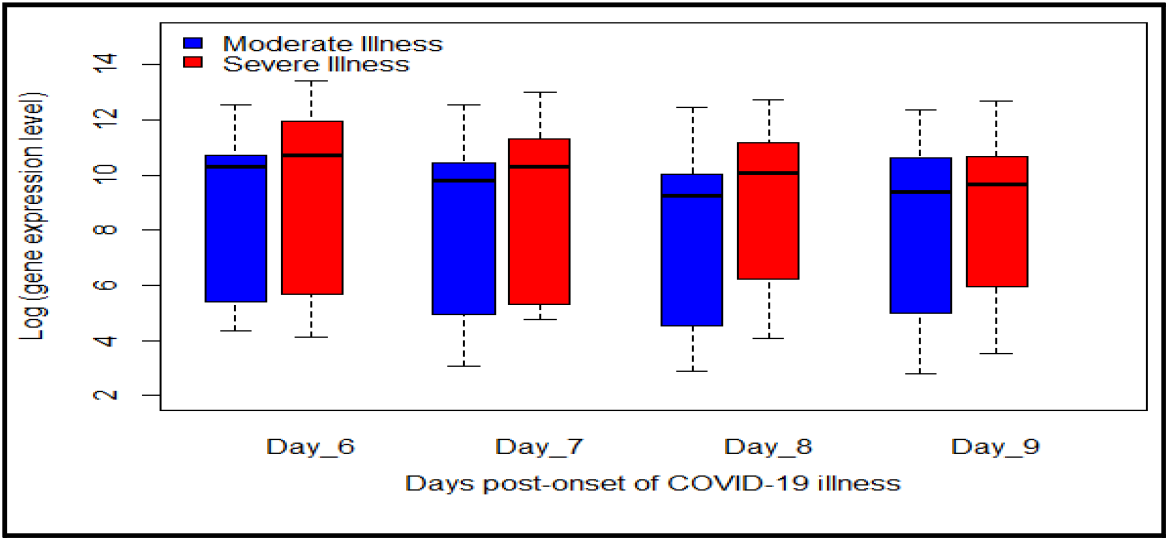
For each of the days 6, 7, 8 and 9 there is higher level of gene expression of NETosis in severe COVID-19 illness compared to moderate illness. Data are from two different patients: one with moderate illness (shown in blue), the other with severe illness (shown in red) sampled at 6, 7, 8, 9 days after onset of illness. For each day, the difference is statistically significant (day 6: p = 0.057; day 7: p = 0.033; day 8: p = 0.015; day 9: p = 0.080). Greatest magnitude of over-expression is observed for the following genes: CEACAM8, CR1, ITGAM and CTSG (2-fold or greater up-regulation in severe illness in at least 3 of the 4 days shown here). Gene expression data were extracted from [13].

### Up-regulation of genes functional in NETosis in lung of COVID-19 patients

NETosis up-regulation was validated in lung tissue from deceased COVID-19 patients. Genes functional in NETosis are up-regulated in the lungs of COVID-19 patients compared to healthy control (Figure 3). These genes include toll-like receptors, cathepsin G, CEA cell adhesion molecule 8, complement C3b/C4b receptor 1, integrin subunit beta 2, C-C motif chemokine ligand 5 and complement factor properdin.

**Figure 3:**
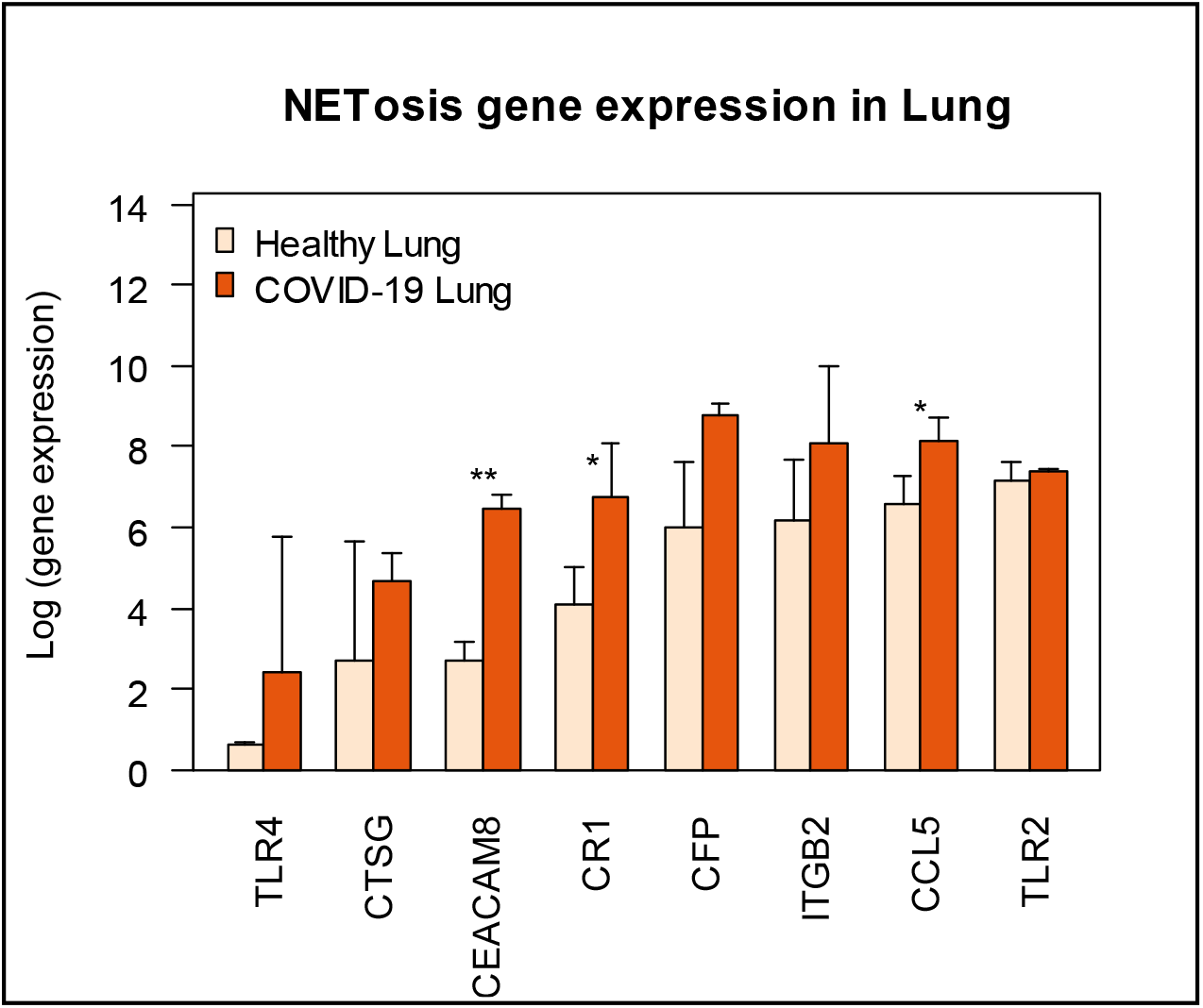
Each bar represents the average log-gene expression in the lung of COVID-19 non-survivors (COVID-19 Lung; n=2) and post-mortem lung tissue of uninfected individuals (Healthy Lung; n=2) [15]. Error bar represents standard deviation of log-gene expression. Significance of up-regulation in severe illness was assessed by t-test and is indicated with an asterisk over the bars (* p < 0.1; ** p < 0.01). The genes functional in NETosis are up-regulated in the lungs of the patients with COVID-19.

### Up-regulation of NETosis is associated with higher neutrophil to lymphocyte ratio (NLR)

NETosis up-regulation was associated with increased NLR in the blood. As shown in Figure 4A, there is up-regulation of genes functional in NETosis at a level higher in the severe case (red) compared to the moderate cases (blue). Additionally, NLR is higher in the blood of severe case compared to moderate cases. With time, both NLR and NETosis gene expression return to baseline in the severe case (red boxes and red bars respectively for case 1 in Figure 4A and 4B).

**Figure 4:**
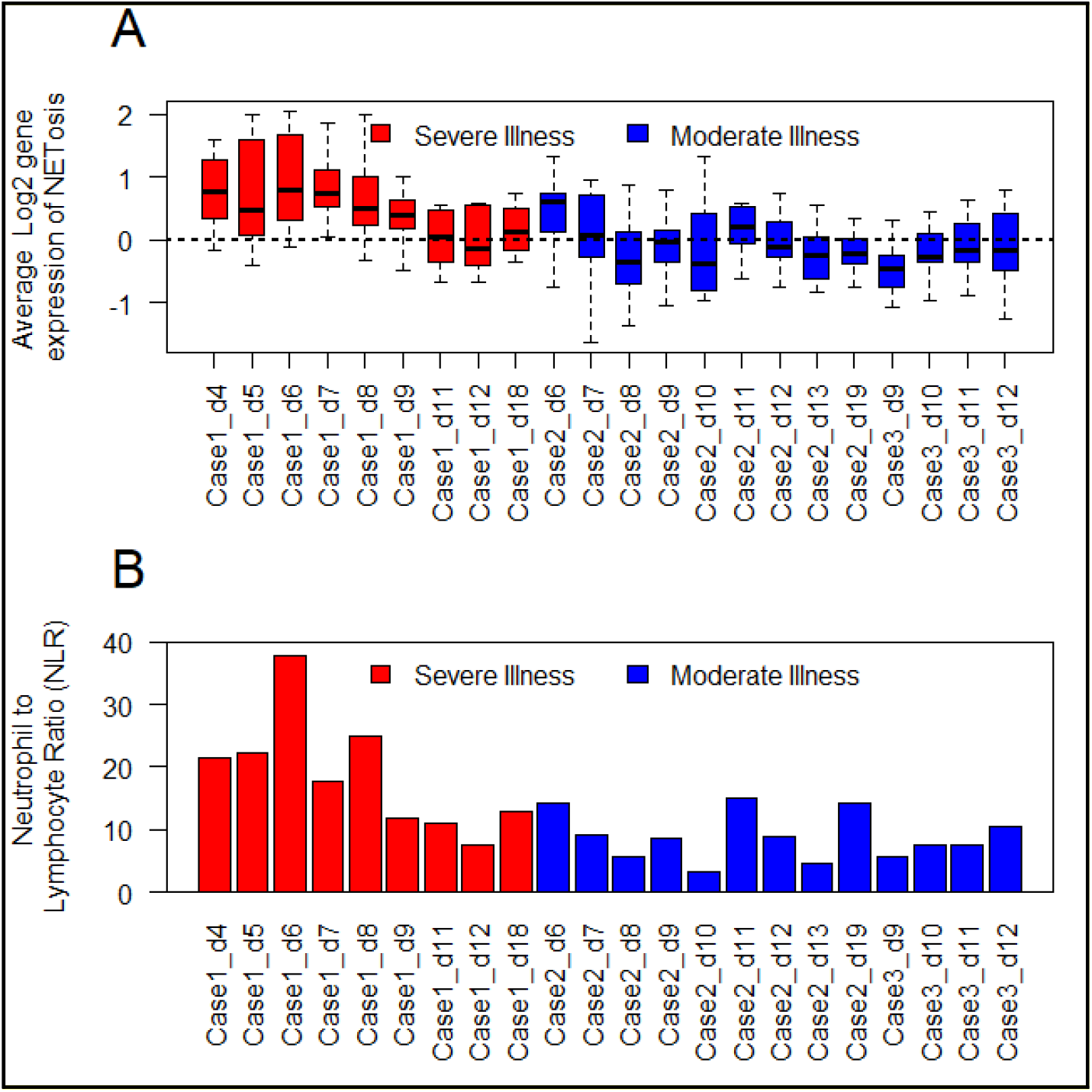
Gene expression data were extracted from [13]. (A) Each box represents a single sample (color-coded red for the patient with severe illness and blue for the patients with moderate illness). Dotted line represents baseline expression (average gene expression in healthy subjects). The samples from the severe case, especially the early time points, have higher expression (up-regulation) of NETosis genes compared to the cases with moderate illness. With passing days from the onset of COVID-19, gene expression tends to return to baseline. (B) Each bar represents neutrophil to lymphocyte ratio (NLR) in peripheral blood in that sample. NLR is increased at the early time points in severe illness. With passing days from the onset of COVID-19, NLR is reduced. Generally, there is agreement in the trend of NETosis gene expression and NLR in the patients of COVID-19 (correlation coefficient = 0.8).

### Negative association between NETosis and respiratory function

In the severely ill patient of COVID-19 line plot was drawn to show the reciprocal relationship of gene expression with oxygen saturation (%). As shown in Figure 5, there is up-regulation of genes (functional in NETosis) at the time of low oxygen saturation and down-regulation otherwise. This is also proven by negative correlation coefficient for the genes (CR1: −0.52, CCL5: −0.37, CFB: −0.57, CFP: −0.12, TLR2: - 0.61, TLR4: −0.65, ITGAM: −0.61, ITGB2: −0.56).

**Figure 5:**
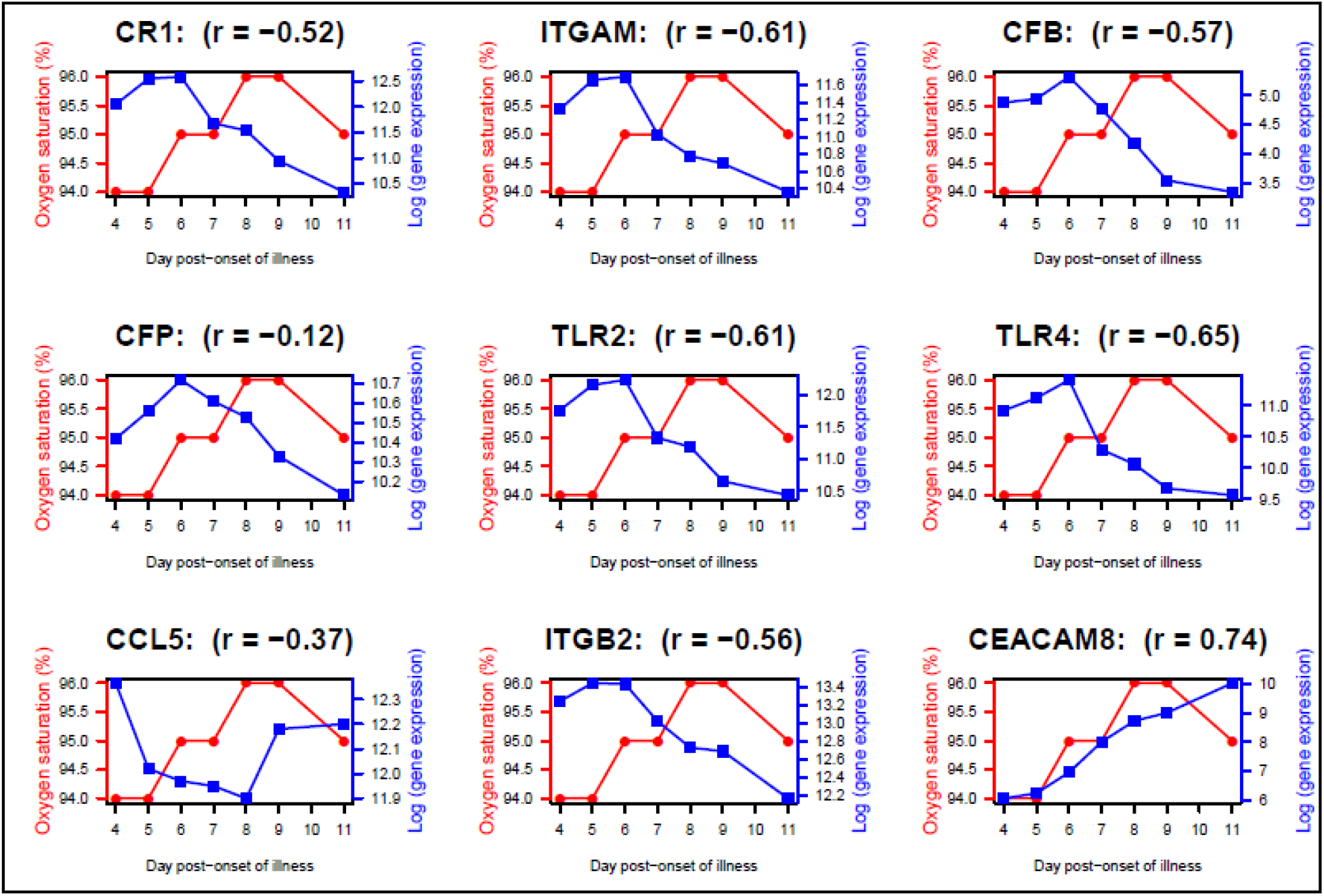
Temporal line plots of selected NETosis genes in the severe patient (Case 1) over days of illness. Each plot corresponds to one gene, with the red line representing oxygen saturation (%), and the blue line representing level of gene expression. Pearson correlation coefficient (r) is negative between oxygen saturation and level of expression of these genes. In contrast to other genes shown here, CEACAM8 up-regulation is sustained over time. Gene expression data were extracted from [13].

### Up-regulation of Complement pathway

Genes belonging to the complement pathway were extracted and subjected to gene set enrichment analysis (permutation testing). As shown in Figure 6, there is significant up-regulation of the genes functional in the complement pathway. Additionally, multiple genes functional in the coagulation cascade are also observed to be up-regulated (Figure 7).

**Figure 6:**
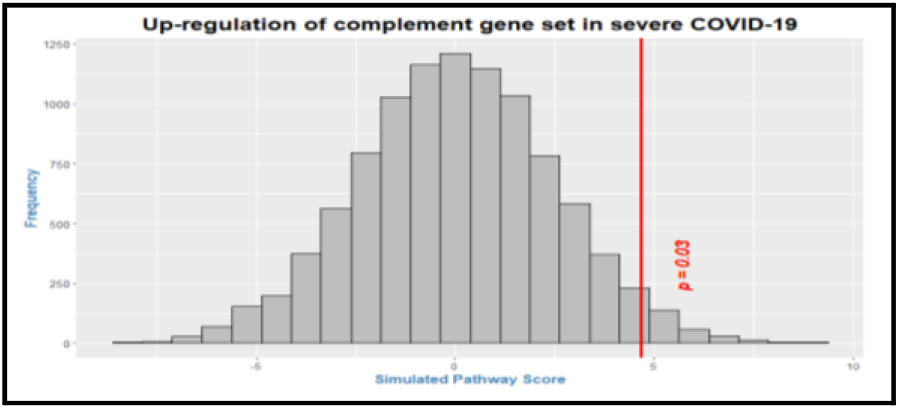
The histogram (gray bars) represents the null distribution of pathway score calculated by weighted averaging of individual relative expression levels (in severe illness with respect to that in moderate illness) for genes functional in the complement pathway. Only those genes of the pathway present on this platform [13] were included in the analysis. The red vertical line represents the observed pathway score for each set. The position of the red line with respect to the histogram (i.e., toward the right tail) suggests that there is significant up-regulation of gene set functional in complement pathway (p = 0.03) in severe COVID-19 illness.

**Figure 7:**
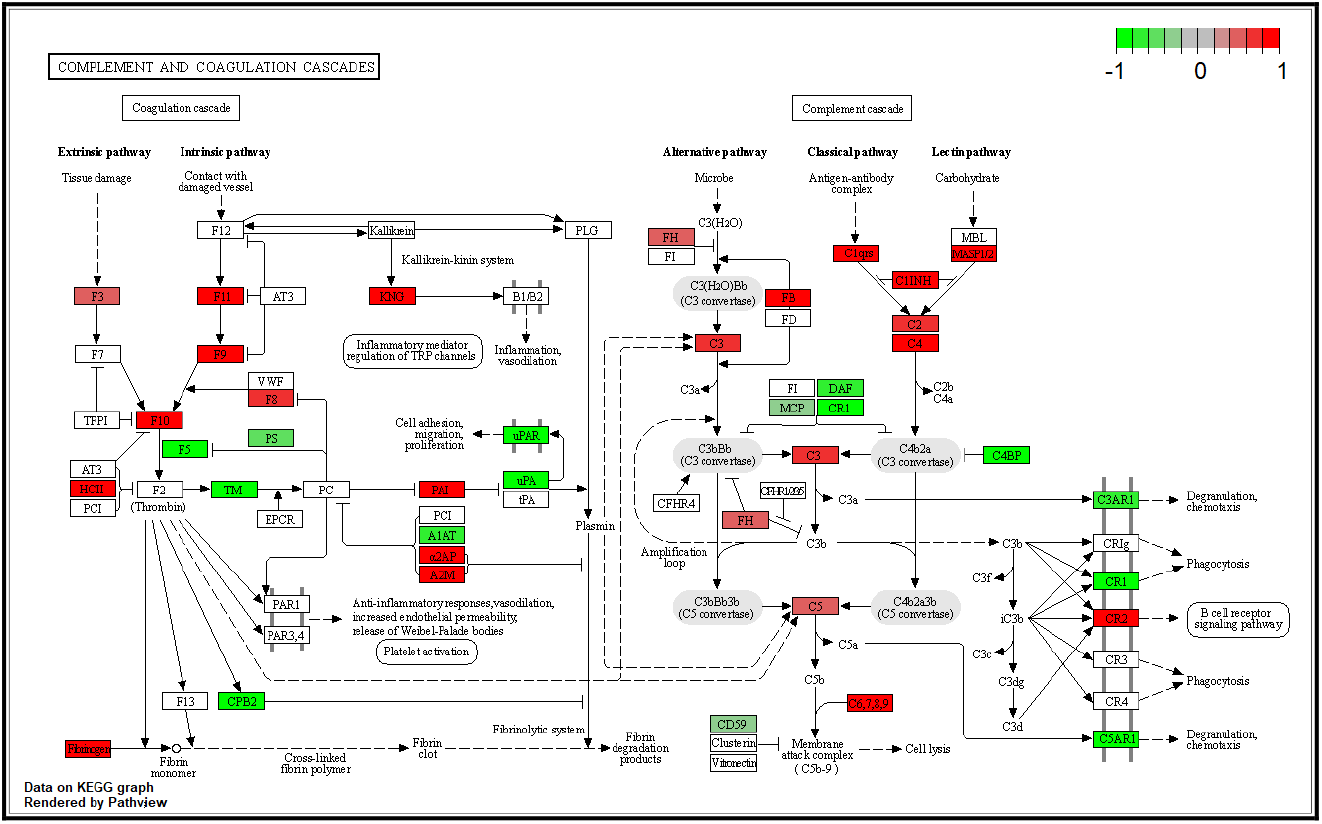
KEGG Complement and Coagulation pathway has been coloured on the basis of differentially expressed genes in COVID-19 using data from [4]. The up-regulated genes are shown in red. The down-regulated genes are shown in green. As displayed, multiple genes of the complement and coagulation cascades are up-regulated in COVID-19.

### IL-6 segregates with complements

We performed an unsupervised clustering of four cytokines (IL-6, IL-8, TNFα, IL1β) with genes functional in NETosis (including complement genes). Gene expression data (log-scale) of all-time samples from the three patients of COVID-19 were used. Hierarchical clustering reveals segregation of IL-6 with complement factors C3, C5 and CFB (Figure 8). The other 3 cytokines form a separate clade. Since the data are captured from peripheral blood of the patients during the time-course of illness, this result suggests role of IL-6 in the gene expression dynamics of complement genes.

**Figure 8:**
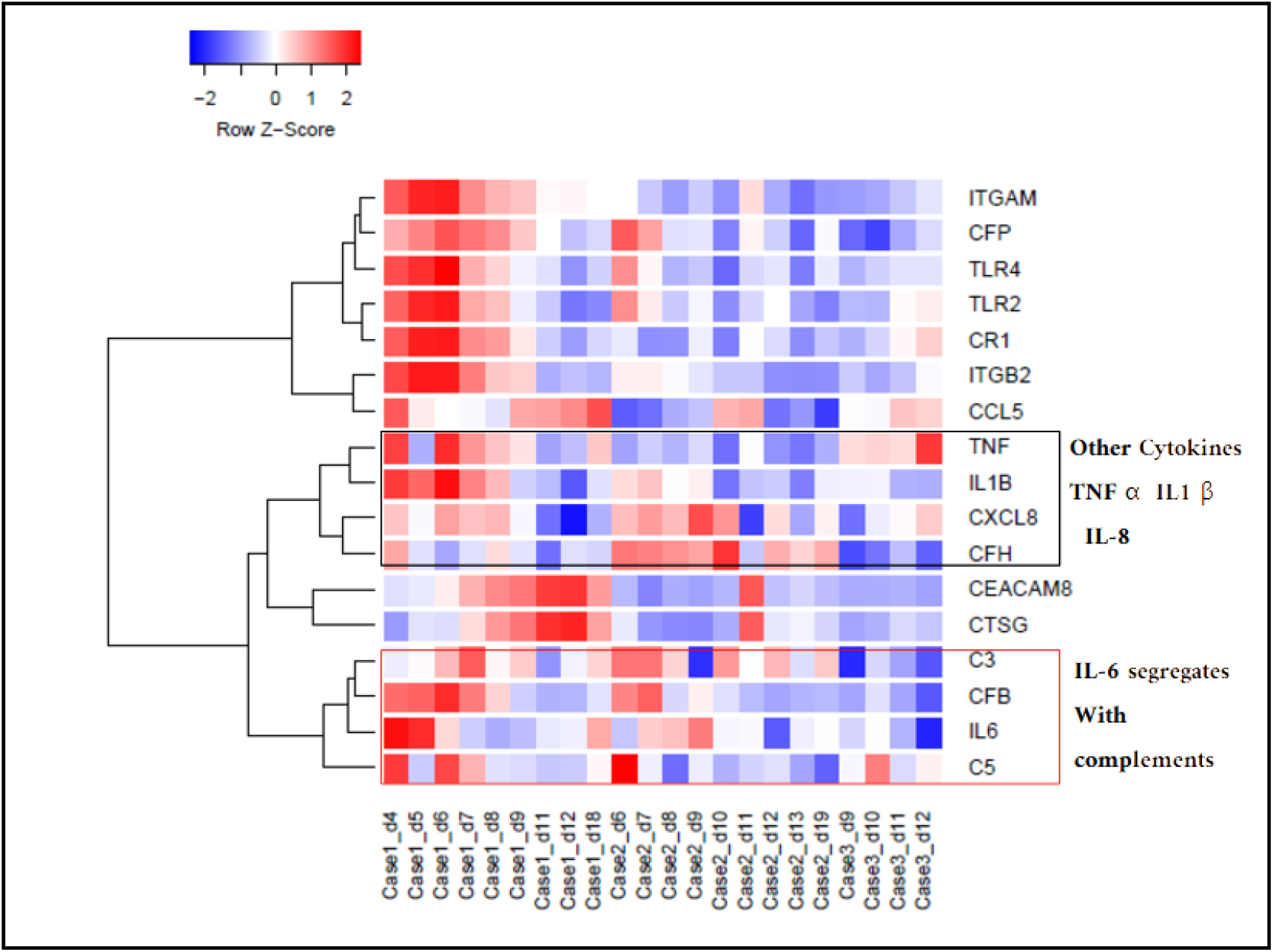
The heat map was generated from all-time samples of the 3 cases of COVID-19 (Case 1 - severe illness, Case 2, 3 - moderate illness) and log (gene expression) of four cytokine genes (IL-6, IL-8, TNFα, IL1β) and genes functional in NETosis (including complement factors) [13]. Unsupervised hierarchical clustering reveals segregation of IL-6 with the complement factors C3, C5 and CFB while the other 3 cytokines (IL-8, TNFα, IL1β) form a separate clade. The samples are arranged according to the case IDs (from left to right: Case 1, Case 2, Case 3), with increasing days post-onset of illness from left to right. Gene expression data were extracted from [13].

### Changing expression level of IL-6

Box plot of longitudinal IL-6 profiling in three groups of subjects (healthy control, moderate illness, severe illness) revealed that the magnitude of up-regulation is greatest early in the disease process [Figure 9]. With increasing days post-onset, the level of expression in the severe illness approaches that in the moderate illness [16]. The reduction in IL-6 expression coincides with increase in the expression of genes functional in NETosis, such as CTSG and CEACAM8 (Figure 10).

**Figure 9:**
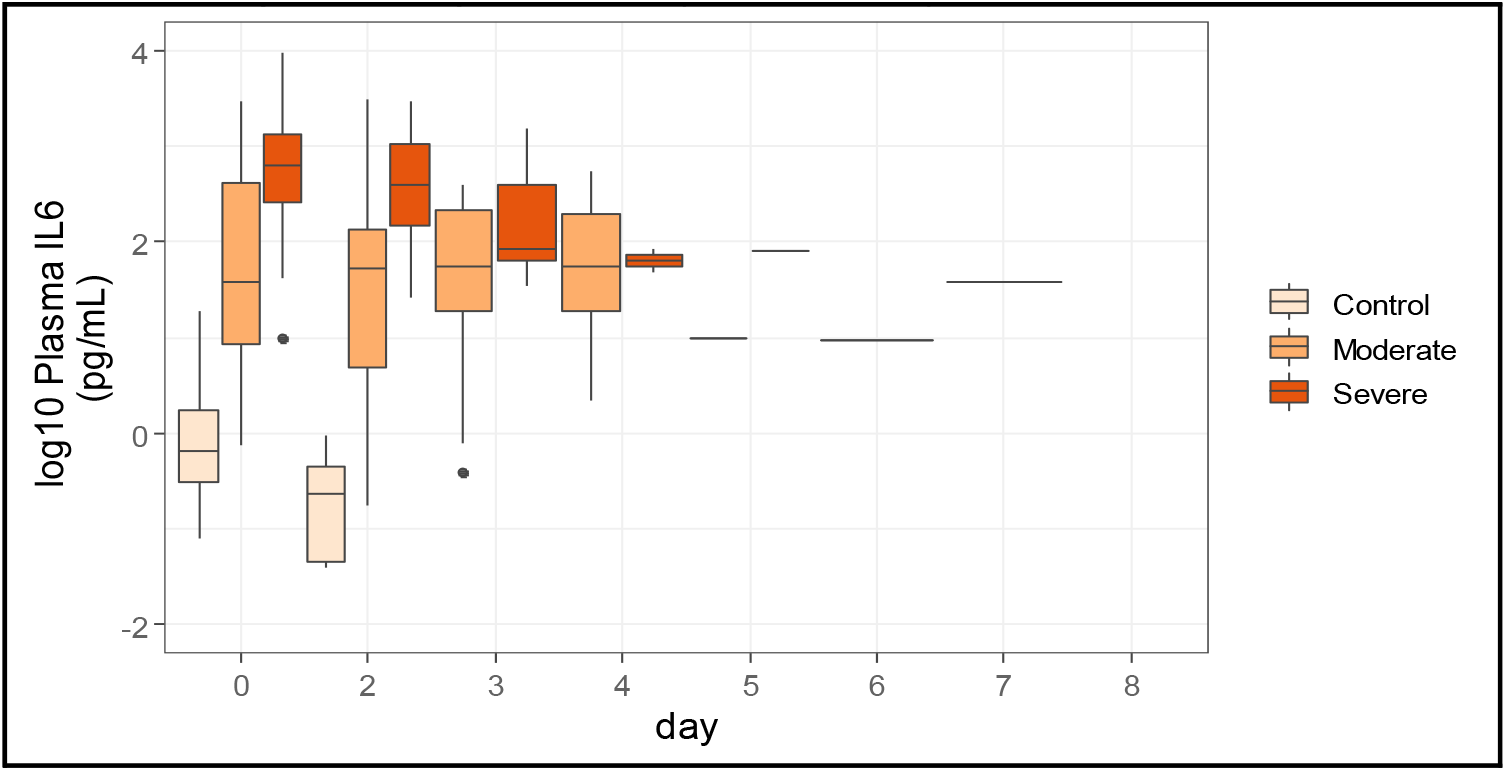
Box plot shows temporal change in plasma IL-6 level in control and patients with (moderate and severe) COVID-19 illness. At the early time points, IL-6 level is higher in the cases with severe illness compared to the cases with moderate illness. At the later time points, the level of IL-6 approaches that in the cases with moderate illness. Data were extracted from [16].

**Figure 10:**
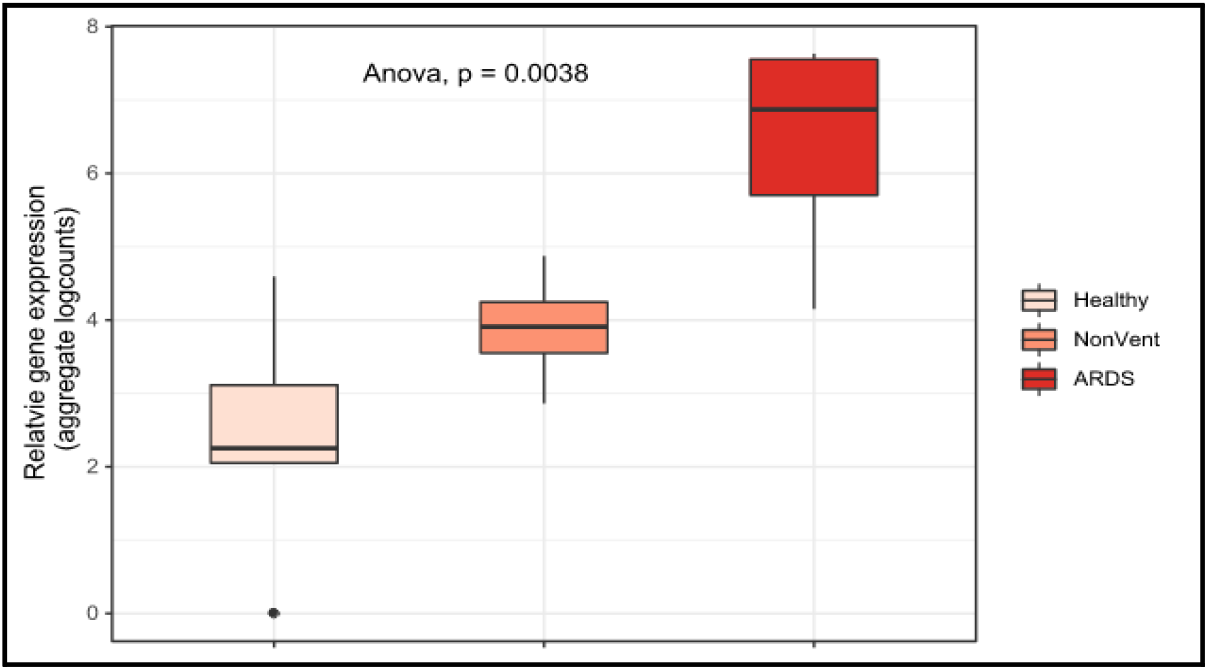
CEACAM8 gene expression in PBMC of healthy control (n=6) and COVID-19 (n=7) were extracted from published data set [14]. Group-level expression (aggregate log counts) data are shown in the box plot, with monotonic up-regulation of CEACAM8 genes from the control group, to COVID-19 cases without ARDS (NonVent) and with ARDS. CEACAM8 up-regulation is a signature of immature or developing neutrophil, a neutrophil subtype associated with COVID-19

### Reduction in the level of DNASE1 expression

In view of the role of DNASE1 in clearance of NETs, we explored the level of its expression in the patients of COVID-19 with varying illness severity. As shown in Figure 11, there is significant down-regulation of DNASE1 in the COVID-19 patients, with greater down-regulation in the patients with ARDS.

**Figure 11:**
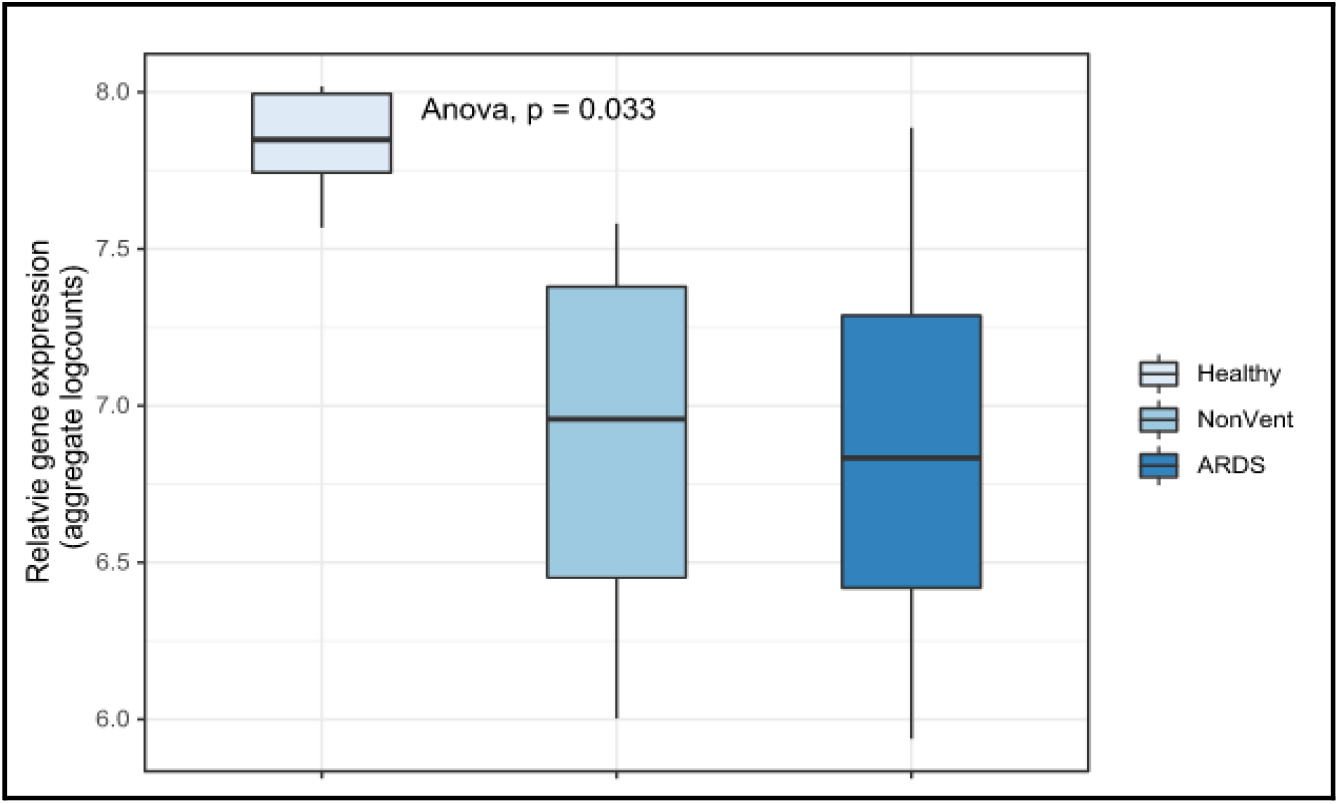
DNASE1 gene expression in PBMC of healthy control (n=6) and COVID-19 (n=7) were extracted from published data set [14]. Box plot of group-level gene expression (aggregate log counts) of DNASE1 shows monotonic down-regulation with increasing severity of illness in COVID-19 cases.

### A model connecting the rise in IL-6 level with NETosis leading to immunothrombosis and ARDS

Considering the robust up-regulation of NETosis, complement pathways and transient early rise of IL-6 in severe cases of COVID-19, a model is proposed. IL-6 is a likely trigger for complement up-regulation and NETosis that leads to coagulation, platelet activation and positive feedback on neutrophil activation. The model was built based on the known effect of IL-6 on up-regulation and activation of complement genes C3 and C5, activation of neutrophil, formation of neutrophil extracellular trap, activation of platelets, coagulation, and positive feedback from platelet to neutrophil. A vicious cycle of NETosis and thrombosis ensues and sustains illness severity and ARDS.

## Discussion

Unbiased analysis of transcriptome data reveals that gene set functional in NETosis is strongly up-regulated in the blood of COVID-19 patients. The up-regulation is statistically significant and is higher in magnitude in severe illness than in moderate illness. Up-regulation of NETosis is most prominent in the early time points of illness, and with passing days, the level of gene expression approaches that in moderate illness. Paired testing reveals that from day 6 until day 9, there is significant and sustained elevation of gene expression functional in NETosis in severe illness compared to moderate illness.

Of note, death of the patients of COVID-19 occurs primarily due to the complications arising from SARS-CoV-2-associated acute respiratory distress syndrome. NETosis is known to cause immunothrombosis and respiratory dysfunction in COVID-19 [5]. We present transcriptional evidence of NETosis up-regulation in both peripheral blood and autopsied lung tissue of COVID-19 patients. Additionally, time-course expression data from a case of severe COVID-19 reveal negative association of NETosis gene expression with respiratory function (oxygen saturation). Together, these findings are consistent with NETosis as an underlying mechanism for a pro-thrombotic state in blood leading to respiratory dysfunction.

Transcriptional profiling of nasopharyngeal swabs from COVID-19 patients have demonstrated upregulation of complement and coagulation pathway associated with mortality and morbidity [4]. Our analysis (of data from [13]) also revealed significant up-regulation of the complement pathway genes in severe COVID-19 illness. NETs act as scaffold for both thrombogenesis and complement activation; and the three pathways (NETosis, complement and coagulation) are considered a single coordinated biological process [17]. NETosing neutrophils have been shown to activate complement via alternative and non-alternative pathways [18]. Also, activated macrophages are known to cause induction of complement factors. The supernatant of macrophage that causes overexpression of the complement factors C3 and CFB are enriched in IL-6 [19], which is consistent with our observation of segregation of complement factors C3 and CFB with IL-6 but not with the other pro-inflammatory cytokines (IL-8, IL-1β and TNFα) in COVID-19 illness. While cooperation among different components of NETosis-complement-coagulation consortium protects host against both haemorrhage and infection [17], unchecked NETosis causes immunothrombosis and leads to acute respiratory distress in COVID-19 illness [5].

By unsupervised clustering, we observed segregation of IL6 with the complement factors C3 and CFB, which are also up-regulated in the target tissue by IL-6-rich supernatant from activated macrophages [19]. Plasma from both COVID-19 patients [5] and patients of sickle cell disease (SCD) with vaso-occlusive crisis (but not from steady state plasma of SCD) cause significant increase in NETosis [20]. The level of IL-6 is observed to be high in the plasma from these patients. Up-regulation of IL-6 signaling has been observed in nasopharyngeal swab [4], lung [15] and has been associated with poor outcome of COVID-19 [2, 4, 15]. Interestingly, Mann and colleagues [7] observed early rise of IL-6 level in critically ill patients of COVID-19, which progressively decreased over time even if the patient did not survive. In data from a different cohort, we also observed an early spike of blood expression of IL-6 in severe COVID-19 illness, which returns, over time, to levels comparable with moderate illness [16]. Together these findings support a dynamic shift in IL-6 level in severe illness – with potential mechanistic and therapeutic significance of IL-6 in the early time window.

It seems likely that the spike in IL-6, secreted by the macrophages responding to the viral entry, triggers NETosis in the patients with severe COVID-19, leading to a complex interaction among NETosis, complement and coagulation pathways [17], pulmonary immunothrombosis and acute respiratory distress. IL-6 is known to stimulate thrombosis in platelet-dependent and platelet-independent manner [5, 21]. With time, while IL-6 levels in patients with severe illness approach that of patients with moderate illness (Figure 8), NETosis and complement activation are sustained [17]. Therefore, inhibition of IL-6 signaling is most beneficial before sustained up-regulation of NETosis by a positive feedback loop (Figure 12). In the later phase, IL-6 levels are similar in severe and moderate illness, but NETosis-complement-coagulation lead to immunothrombosis and acute respiratory distress in the severe cases. In this phase, a different strategy is called for, such as, inhibitor of complement and NETosis. Level of IL-6 upon admission can be used as a prognostic marker of outcome [2] and for prioritization of anti-IL-6 therapy.

**Figure 12:**
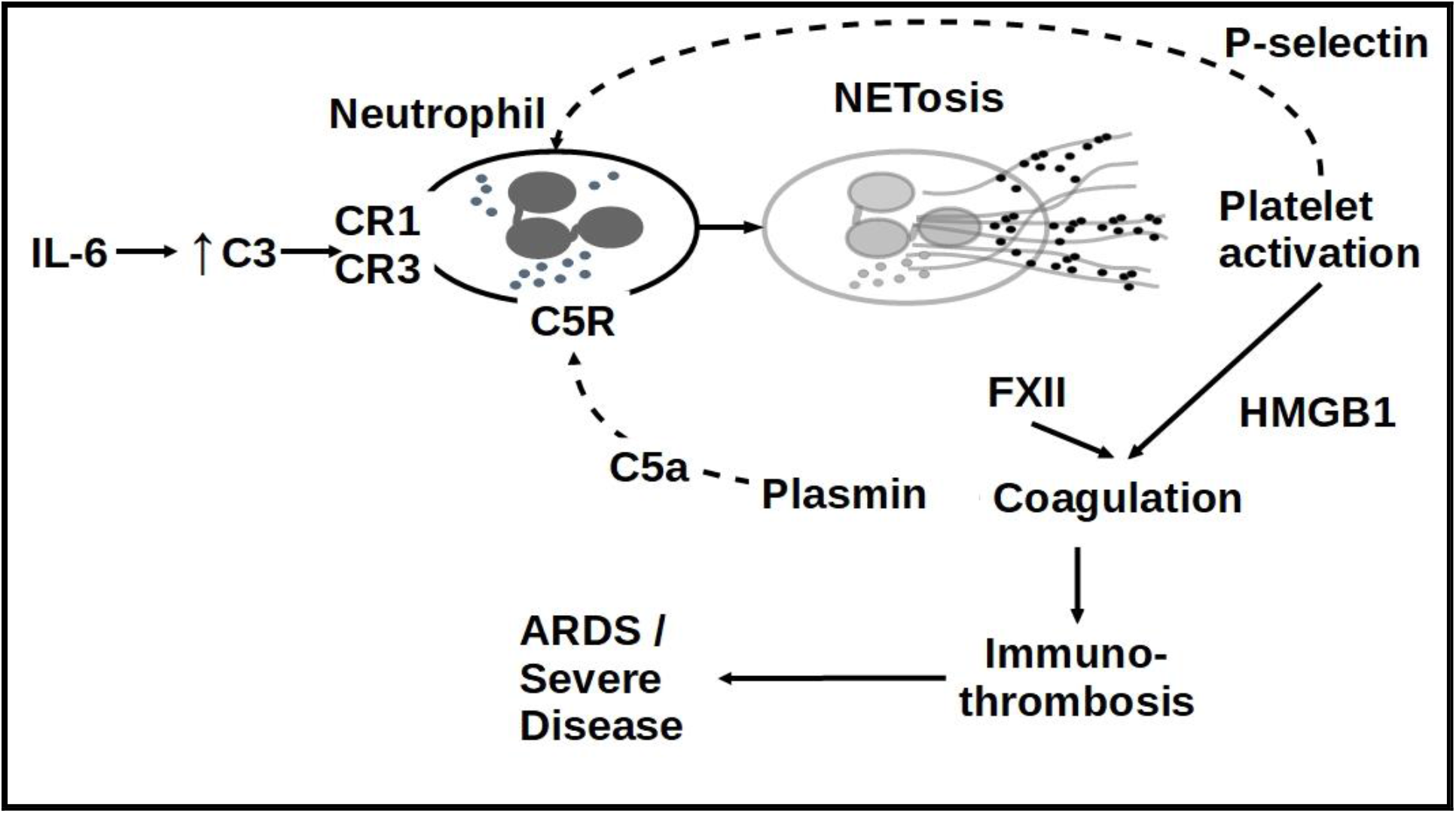
NETosis, complement activation and coagulation are functionally inter-related and together produce immuno-thrombosis. As the blood vessels in the lungs are clogged, it leads to acute respiratory distress and high mortality. IL-6 induces expression of C3 and CFB leading to complement activation. C3 engages complement receptors (CR1, CR3) on neutrophil and activates formation of NET [17]. The components of NET activate platelets which secrete HMGB1, inducing coagulation. Negatively charged NETs can bind and activate circulating glycoprotein FXII (a zymogen produced by liver) that induce coagulation. Plasmin cleaves C5 to active C5a which in turn activates Neutrophil by binding via C5R [17]. Activated platelets can engage with activated neutrophils through binding of P-selectin to PSGL-1[17].

Neutrophil to lymphocyte ratio (NLR) is proposed as a prognostic biomarker of disease severity and organ failure in COVID-19 [6]. In general, there is an increased number of neutrophils in blood, which, along with lymphopenia, contribute to high NLR. There is also increased neutrophil activation of genes functional in formation of NET. The gene expression in COVID-19 is consistent with neutrophilia commonly observed in severe COVID-19 illness resulting in increased formation of Neutrophil Extracellular Traps (NETs). Therefore, NETosis adequately explains the prognostic power of NLR, and extends itself as a fundamental dysregulation underlying COVID-19 disease severity, respiratory distress and mortality.

While there is increased number of neutrophil in the patients of COVID-19, it is not clear if these are the usual neutrophils of healthy blood. Wilk and colleagues [14] observed a novel kind of “developing neutrophil” in the blood of COVID-19 patients. These neutrophils express high level of CEACAM8, a marker of immature neutrophils that are higher in men and pregnant women compared to non-pregnant women [22]. Notably, mortality and morbidity in COVID-19 has been consistently associated with gender of the patients, with male patients at a higher risk of poor outcome [3, 23]. The role of any hormonal influence on neutrophil type and activation (and NETosis) in COVID-19 outcome remains to be elucidated.

Sepsis appears as the single most frequent factor associated with mortality in COVID-19 [24]. Similar to COVID-19, sepsis is also associated with coagulopathy [25, 26]. It is likely that the dynamic cytokine (IL-6) dysregulation induces NETosis and coagulation in other non-COVID causes of sepsis. Thus, temporal and precise mechanistic therapy targeting IL-6 and NETosis shall benefit critically ill patients of both COVID-19 and sepsis.

## Conclusion

In this study, we present robust transcriptomic evidence of NETosis and complement activation in COVID-19. Level of IL-6 expression fluctuates, with a spike at the beginning that triggers NETosis in the severe cases of COVID-19. Therefore, anti-IL-6 therapy should be initiated as early as possible in severe cases. However, once the patient has entered a state of sustained NETosis-complement activation and immunothrombosis, blocking IL-6 signaling needs to be supplemented with additional measures such as inhibition of NETosis or complement pathway. Timing matters for initiation of the right therapy, and for the right patient.

## Methods

### Selection and preprocessing of the data

Gene expression data of different covid-19 studies were downloaded from NCBI GEO [27] and ArrayExpress[28] data repository portals (Table 1). The raw count matrix data were quantile normalised and log transformed where necessary before analysis. The normalised data were then stored as individual “expressionSet” objects and subjected to downstream analysis. All analyses were performed in the R programming language [29].

**Table 1:** Study characteristics with sources for data sets used in the current study.Code and data availability. Data and R code is available in github repository named **covid-19** (“https://github.com/skm-lab/covid-19”).

| Database Link | Author | Article Title | DOI | Country | Tissue | Ref. | Used in the Figure of manuscript |
| --- | --- | --- | --- | --- | --- | --- | --- |
| https://covidgenes.weill.cornell.edu/ | Ramlall | Immune complement and coagulation dysfunction in adverse outcomes of SARS-CoV-2 infection | 10.1038/s41591-020-1021-2 | USA | Nasopharyngeal Swabs | 4 | 12 |
| E-MTAB-8871 (ArrayExpress) | Ong | A Dynamic Immune Response Shapes COVID-19 Progression | 10.1016/j.chom.2020.03.021 | China | Whole blood | 13 | 1, 2, 4-7 |
| GSE150728 (NCBI GEO) | Wilk | A single-cell atlas of the peripheral immune response in patients with severe COVID-19 | 10.1038/s41591-020-0944-y | USA | PBMC | 14 | 1, 9, 14 |
| GSE147507 (NCBI GEO) | Blanco-Melo | SARS-CoV-2 launches a unique transcriptional signature from in vitro, ex vivo, and in vivo systems | 10.1101/2020.03.24.004655 | USA | Primary cell cultered from Lung | 17 | 3 |
| https://www.immport.org/shared/home; study ID SDY1655 | Lucas | Longitudinal analyses reveal immunological misfiring in severe COVID-19 | 10.1038/s41586-020-2588-y | USA | Serum | 18 | 8 |

### Permutation based gene set enrichment analysis

Any pathway with 10 or less number of genes was discarded from analysis. For each gene, t-statistic was computed to denote change in gene expression in case group compared to the control group. For each pathway, a score was calculated by weighted averaging (i.e., sum of the gene-level t-statistics divided by the square root of the number of genes in the pathway) of all gene-level t-statistics for the pathway. Significance of the observed pathway score was calculated by permutation testing performed in the following manner. In each permutation, the samples were randomly re-labelled as case and control, with calculation of a simulated pathway score. This was done 10,000 times generating 10,000 simulated values representing the null distribution of the pathway score. Deviation of the observed pathway score from the null distribution was quantified by the fraction of times that the simulated score was more extreme than the observed score. This result was assigned as permutation p-value of the observed pathway score. Pathway enrichment analysis was performed using code modified from the R function gseattperm() of the package Category [30].

### Selection of cytokine and NETosis genes

The genes belonging to two groups were selected from relevant literature reviews on cytokine storm [31] and NETosis[17].

### Method for Deconvolution

We used the CIBERSORTx [32] for deconvolution of transcriptome data i.e., to find the cellular components in the sample through search for similarity of expression with reference expression values of specific cell types. A gene by sample expression matrix was created with the instructed format of the web portal guidelines. The reference immune cell gene expression was selected from the leukocyte signature matrix (LM22)[33]. The analysis was run without batch correction (only one dataset at a time) and normalisation (as instructed for RNAseq data) with 1000 permutations. The resulting sample by immune cell-fraction matrix was downloaded in comma separated values (.csv) file format and analysed to estimate neutrophil to lymphocyte ratio (NLR).

## Data Availability

Data and R code is available in github repository named covid-19 (https://github.com/skm-lab/covid-19).

## Code and data availability

Data and R code is available in github repository named **covid-19** (“https://github.com/skm-lab/covid-19”).

## Acknowledgement

SKM acknowledges helpful input on platelet-neutrophil interaction from Prof. Debabrata Dash, Department of Biochemistry, Institute of Medical Sciences, Banaras Hindu University, Varanasi.

